# Transmission dynamics of COVID-19 in the outbreak among healthcare workers at a university hospital in Rio de Janeiro, Brazil

**DOI:** 10.1101/2024.11.13.24317283

**Authors:** José Ueleres Braga, José Hérmogennes Rocco Suassuna, Ana Sara Semeão de Souza

## Abstract

**Objective:** The objective of this study was to investigate an outbreak of COVID-19 among HCWs in the nephrology service of a university hospital in Rio de Janeiro in March 2020.

**Methods:** The study population consisted of a team of 59 HCWs, and surveys were conducted through digital interviews. Signs and symptoms, existence of comorbidities, hospitalization, and the network of contacts among the professionals during the period from March 1 to 23, 2020 were evaluated. The analysis consisted of: (i) describe epidemic curve, (ii) transmission chains, (iii) estimate the effective reproduction number (R_t_), and (iv) identification of factors related to infectious cases.

**Results:** Of the 59 professionals in the nephrology service, 43 (73%) participated in the study. The first case occurred on March 1 and the last case occurred on March 23, 2020. The outbreak peaked on March 12–13. 31 participants were probable or confirmed cases COVID-19. These were mostly women (71%), with an average age of 37 years, and were predominantly doctors (45%). Only one participant was hospitalized. The risk factors for contracting COVID-19 were being a medical doctor, working at another hospital, and being female.

**Conclusion:** We concluded that there was an outbreak of COVID-19 among health professionals in the nephrology service at HUPE from March 1 to 23, 2020. The transmission chain network had large clusters showing intense transmission.

## Background

The Coronavirus Disease 2019 (COVID-19) epidemic, caused by severe acute respiratory syndrome coronavirus 2 (SARS-CoV-2), started in Wuhan, China in December 2019 and was initially located only in hospital settings before becoming a public health problem^1^. On January 30, 2020, the World Health Organization (WHO) announced that the epidemic was a public health emergency of international interest^2^.

As of May 14, 2020, WHO reported a total of 4,248,389 confirmed cases and 294,046 deaths from the disease in 215 countries. The United States, Spain, Russia, United Kingdom, Italy, Germany, and Brazil have the highest numbers of cases so far^3^. On February 26, the first case of COVID-19 was confirmed in Brazil, which was also the first case in Latin America^4^.

In the State of Rio de Janeiro, the first probable case was identified on March 2 and was confirmed on the March 6^5^. On March 16, 2020, the press reported a possible outbreak of COVID-19 in “Médicos de hospital universitário do Rio são diagnosticados com coronavírus”^6^. The article indicated that doctors on the university hospital’s nephrology service tested positive for coronavirus, and one was hospitalized in serious condition^6^. It seemed that we were dealing with a nosocomial outbreak of an emerging infectious disease that deserved further investigation.

Since then, extensive measures have been implemented to reduce the transmission of COVID-19 from person to person to control the current outbreak^7,8^. Special attention and efforts to protect against or reduce transmission should be applied to susceptible populations, especially healthcare workers (HCWs), children, and the elderly. The transmission of the virus in hospitals has been reported, and COVID-19 cases have been identified among health professionals^9^. According to Chang et. al.^10^, the WHO confirmed 8,098 cases and 774 (9.6%) deaths during the SARS epidemic in 2002, and healthcare professionals comprised 1,707 (21%) of the cases. The objective of this study was to investigate an outbreak of COVID-19 among HCWs in the nephrology service of a university hospital in Rio de Janeiro in March 2020. The specific objectives were: (i) to characterize the outbreak, regarding the person-place-time triad, (ii) analyze the epidemic curve, (iii) identify the transmission chains, and (iv) estimate the probability of transmission, that is the effective reproduction number, and evaluate the transmissibility of COVID-19.

## Methods

The study population consisted of a team of 59 health professionals in the nephrology service of a university hospital in the city of Rio de Janeiro, Rio de Janeiro, Brazil. Surveys were conducted through digital interviews (electronic mail and electronic forms) and telephone calls to monitor responses and minimize losses.

The signs and symptoms that the professionals experienced during the period from March 1 to 23, 2020 were evaluated. This period was selected based on the knowledge that the symptoms of COVID-19 infection typically appear after an average 5.2 day incubation period^11^.

The study subjects were evaluated daily for the presence of related symptoms, including fever, cough, sore throat, difficulty breathing, myalgia, diarrhea, nausea and vomiting, headache, runny nose, irritability or confusion, altered taste, altered smell, adynamia, and others. During the same period, clinical signs were also evaluated, including fever, pharyngeal exudate, seizure, conjunctivitis, coma, dyspnea or tachypnea, alteration of pulmonary auscultation, and alteration of chest radiology (or other imaging). We also investigated the existence of comorbidities including cardiovascular diseases, such as hypertension; diabetes; liver disease; chronic or neuromuscular neurological disease; immunodeficiency; HIV infection; kidney disease; asthma; and other chronic lung diseases and neoplasia (solid or hematological tumor).

Hospitalization was also assessed during the study period (3/1/2020 to 3/23/2020), including the reason for hospitalization and the dates of admission and discharge. Whether mechanical ventilation was used was also noted. The study participants were also asked about laboratory tests for COVID-19, including the date of sample collection (first and second) and the results. Quarantine (home) was also evaluated, including the start and end dates, and their health on the day that they answered the questionnaire.

The network of contacts among the professionals in the nephrology service was evaluated through registration of contacts with other members of the technical team. Study participants were asked on each day of the study period (3/1/2020 to 3/23/2020) whether the professional was in contact with any of the other workers. The duration of contact was also assessed and placed in one of three categories: ≥60 minutes, <60 minutes, or no contact.

During the study period, the tests for COVID-19 were still limited in Brazil only to severe cases, therefore, those who presented clinical signs and symptoms compatible with COVID-19 and had contact with a confirmed case were considered as probable cases. laboratory tests (RT-PCR) or consistent clinical signs, typical tomographic images, and positive serological tests for the SARS-CoV-2 virus. Confirmed cases showed compatible clinical symptoms and a positive SARS-CoV-2 RT-PCR test result. We constructed transmission chains based on the time of exposure and the appearance of signs, symptoms, and positive laboratory test results. The analysis plan consisted of three stages: (i) classification of cases (index and secondary), (ii) calculation of the R_t_ transmissibility measures for each consequent period from the first identified case to estimate and model variations in the magnitude of transmission, and (iii) identification of factors related to infectious cases (for example, age, sex, signs, and symptoms) and factors related to individual susceptibility (workload, type of work, etc.). Transmission chains were built only among participants who had some contact that represented a risk of transmission. Logistic regression models were used to estimate the odds ratio for COVID-19 infection as the primary outcome, considering each transmission factor and susceptibility factor.

The analyses were performed using the “epicontacts” and “EpiEstim” packages from the R Epidemics Consortium project (https://www.repidemicsconsortium.org/projects/). The research project was approved by the Research Ethics Committee (CEP) of the Social Medicine

Institute (IMS) of the State University of Rio de Janeiro (UERJ) (CAAE: 30685220.8.0000.5260). Besides, written assent was sought from each participant.

## Results

Of the 59 professionals in the nephrology service, 43 (73%) participated in the study. Of these, 67% were female, with an average age of 40 years (range, 24–69 years), 30% reported chronic disease, 70% were doctors, and 63% had another job.

The first case occurred on March 1 and the last case occurred on March 23, 2020. The outbreak peaked on March 12–13. The shape of the epidemic curve is similar to that observed in outbreaks with a few or a single source of infection (Figure 1).

**Figure 1.**
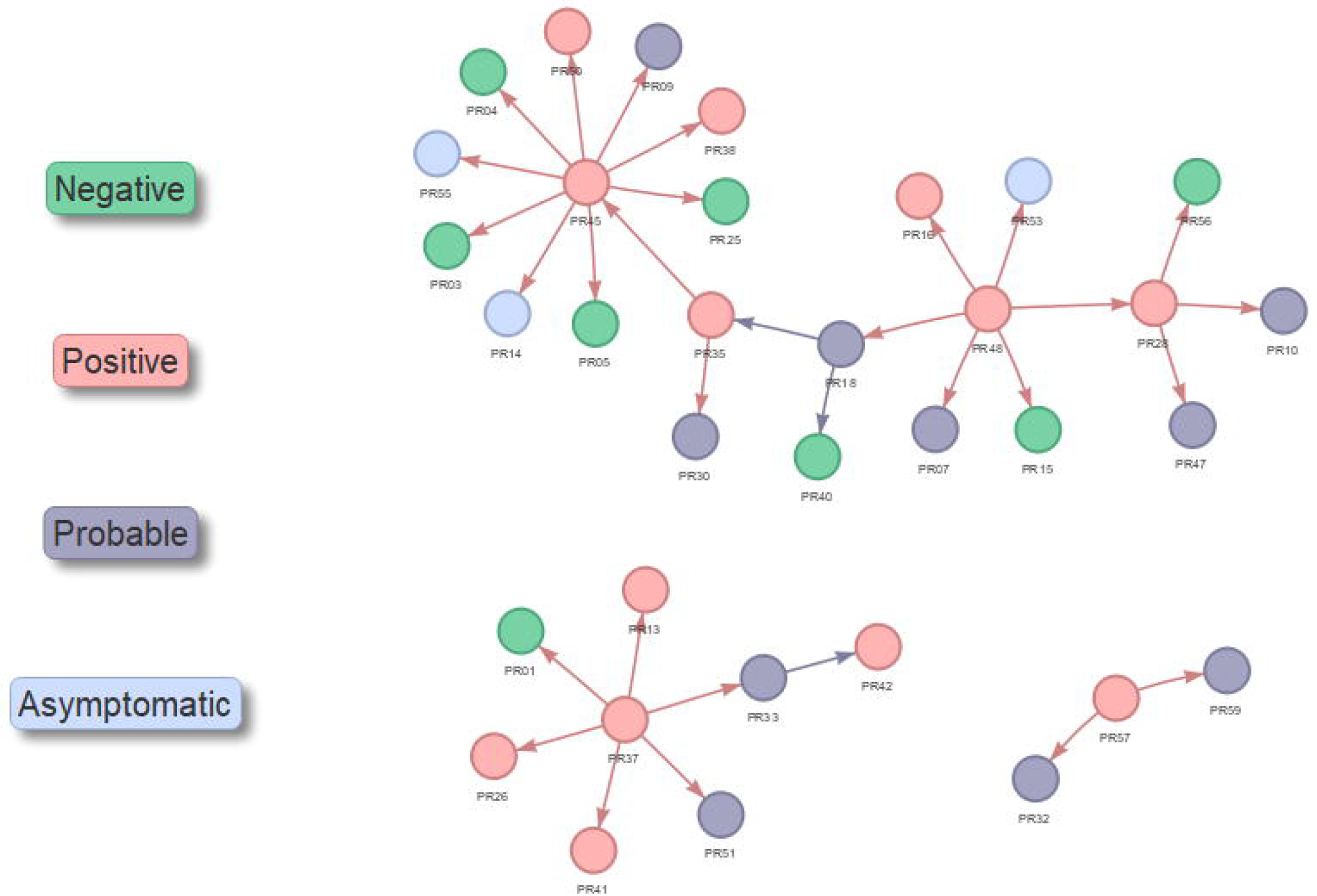
Epidemic curve of probable and confirmed cases between March 1 to March 23, 2020

The chronic diseases reported were cardiovascular diseases, including hypertension (14%), asthma (14%), other chronic lung disease (5%), and neoplastic disease (solid or hematological tumor, 5%). No one had diabetes, liver disease, chronic neurological or neuromuscular diseases, immunodeficiency, HIV infection, or kidney disease. Participants who fell ill with COVID-19 had the following symptoms: adynamia (81%), headache (71%), runny nose (67%), change in taste (67%), change in smell (67%), cough (62%), fever (57%), sore throat (57%), myalgia (52%), diarrhea (43%), difficulty breathing (29%), nausea/vomiting (14%), pharyngeal exudate (14%), dyspnea (14%), change in pulmonary auscultation (10%), conjunctivitis (5%), irritability/confusion (5%), and a change in chest radiology (5%).

Of the 43 participants, 34 (79%) were classified as a probable or confirmed case of COVID-19. Most of these were women (71%), with an average age of 37 years and predominantly doctors (45%). Only one participant was hospitalized.

A visualization of the chains of contact for each COVID-19 case is shown in Figure 2. This chart shows three large clusters of cases, PR45, PR37, and PR48, with 10, 6, and 6 primary contacts, respectively. On average, each case had 1.05 contacts (corresponding to the number of edges or connections to other nodes), which is compatible with the reported transmission of SARS-CoV-2. The number of ranged from 1–10. Examination of the transmission chains according to the participants’ characteristics (e.g., sex, professional category, and activities) did not show obvious signs of non-random mixing patterns.

**Figure 2.**
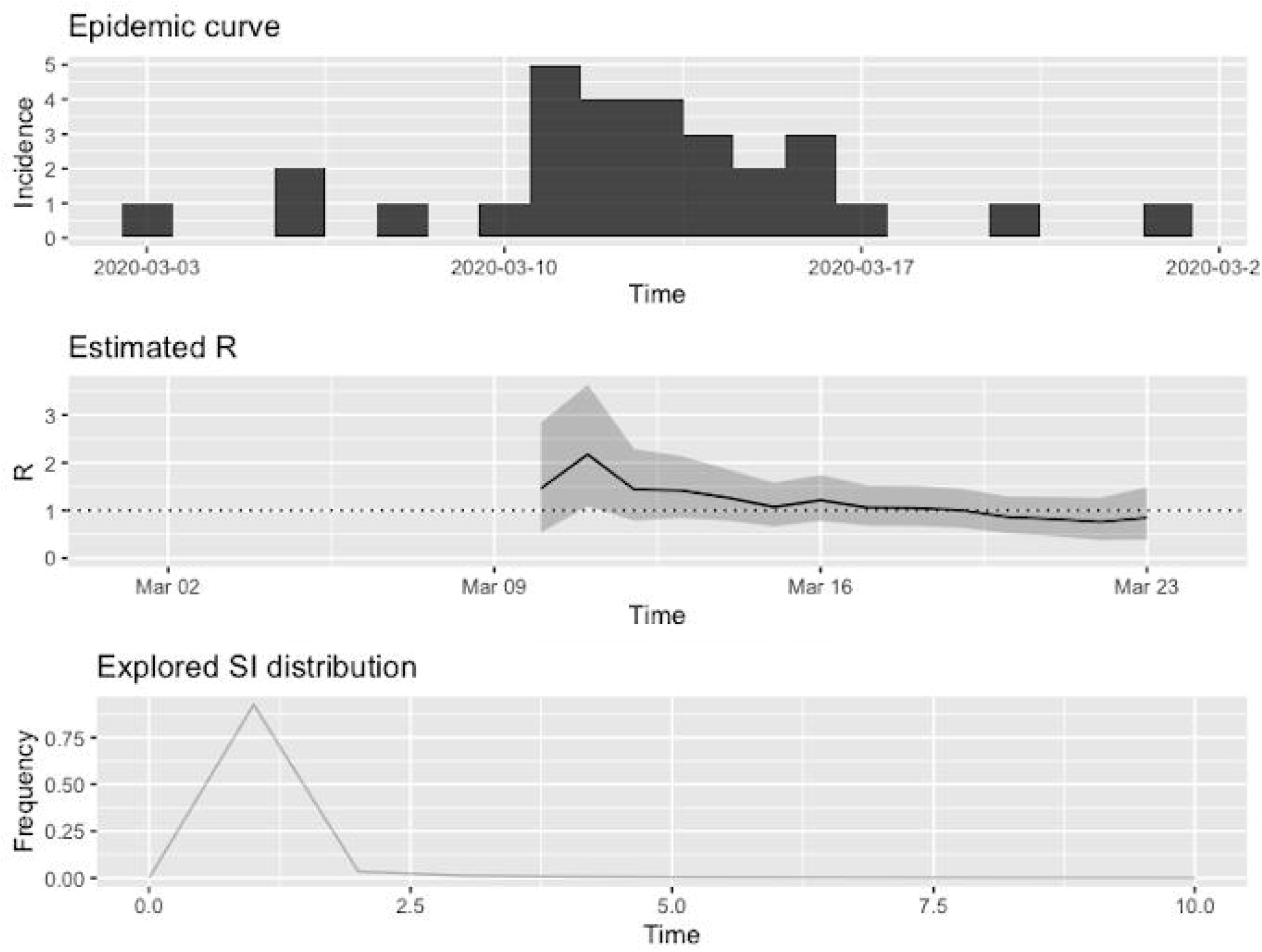
COVID-19 transmission chains generated by contact tracing of cases

The distribution of the serial intervals (i.e., the delay between primary and secondary onset), showed a median of 5.00 days and a mean of 6.62 days. To evaluate SARS-CoV-2 transmission, we generated three graphs, including an epidemic curve of secondary cases. We plotted the epidemic curves by date of onset of COVID-19 symptoms and determined the estimated Rt values and their 95% confidence intervals using non-parametric methods (Figure 3). Finally, the serial interval distribution was plotted, which showed that the interval was predominantly less than two days. The reproduction number (Rt) of COVID-19 substantially decreased after the peak of the curve on Mar 12, 2020 and has remained below 1.

**Figure 3.**
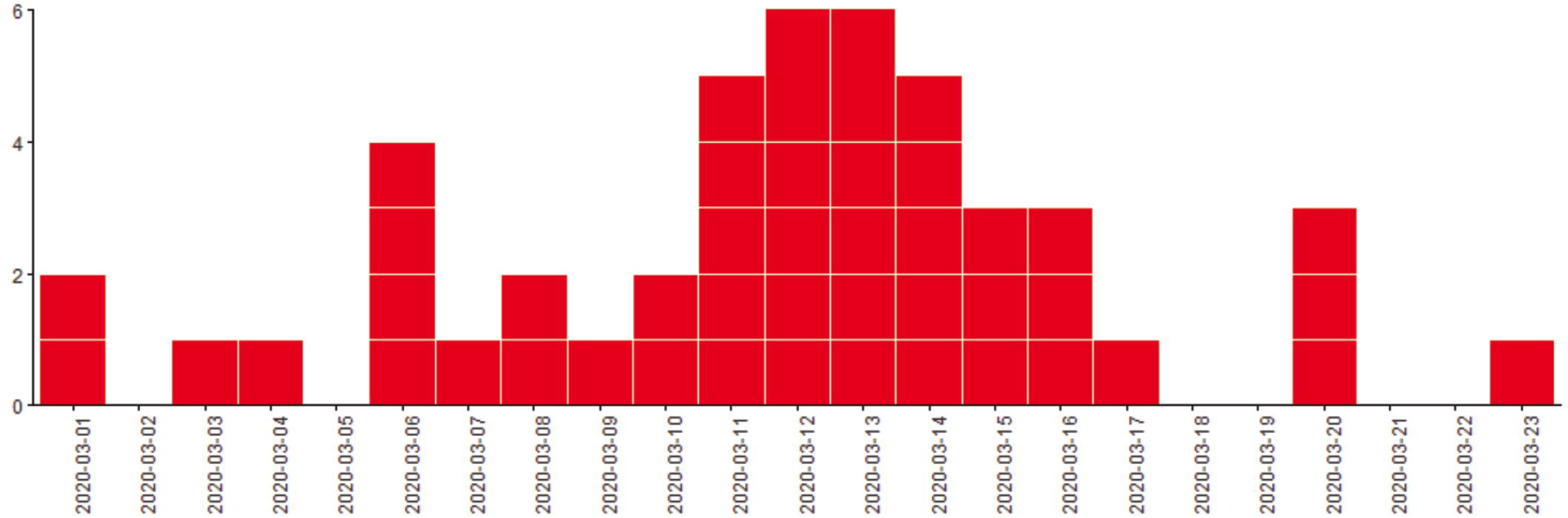
Epidemic curve of secondary cases and the effective reproduction number

The risk factors for SARS-CoV-2 infection during this outbreak were being a medical doctor (OR = 4.76, p=0.053), working in another hospital (OR = 3.13, p=0.115), and being female (OR = 3.03, p=0.173).

## Discussion

Based on the findings of this study, we concluded that there was an outbreak of COVID- 19 among health professionals in the nephrology service at university hospital from March 1–23, 2020. The transmission chain network indicated the presence of large clusters and intense transmission. The main risk factors were being a medical doctor, working in another hospital, and being female. Based on a literature review, this is the first investigation of a COVID-19 outbreak among health professionals, and no other study on hospital outbreak investigation has assessed the intensity and transmission networks.

Nguyen et. al.^12^ conducted a prospective cohort study to examine the risk of COVID-19 among frontline HCWs and concluded that frontline HCWs had a relative risk of 11.6 (95% CI: 10.9–12.3) for a positive test when compared with the general community. It is known that work-related transmission among HCWs constitutes a large proportion of the cases in coronavirus outbreaks. HCWs comprised a large proportion of suspected severe acute respiratory syndrome (SARS) cases in Asia (37–63%) and Middle East respiratory syndrome (MERS) cases (43.5%)^13^. Despite precautions taken against nosocomial transmission, a high prevalence of SARS-CoV-2 infections among HCWs is expected. Lan et. al. 2020^14^ studied work-related transmission in four COVID-19 outbreaks and found that an elevated risk of infection was not limited to HCWs, although COVID-19 cases were the highest among HCWs, comprising 22% of all work-related cases.

Zhou et al.^15^ recently conducted a rapid review and meta-analysis of nosocomial SARS-CoV-2 infections. They reviewed 20 studies that included HCWs, and the results showed that those infected consisted of nurses (56.0%), medical doctors (33.0%), and other staff, such as careers, cleaners, and hospital support staff (11.0%). Finally, a limitation of the study was to consider that all transmission between health professionals took place from another health professional.

## Conclusion

It is difficult for medical doctors to take all the necessary precautions when they come into contact with infected patients, much less when they interact with other members of the healthcare team. These professionals are often grouped around computers and other work equipment. When the professionals are among themselves, they often neglect protective measures against cross-contamination. Our results corroborate this, since the transmission rate in this outbreak was high. Therefore, it serves as a warning. The outbreak was probably established by close contact among the HCWs outside the patient wards.

This study highlights the importance of understanding the transmission of this disease not only in the hospital environment but also among the essential workers who treat critically ill patients requiring hospitalization.

## Data Availability

Participants' data is sensitive and therefore cannot be made available.

## Declarations

## Acknowledgments

We would like to extend our gratitude to the nephrology service of the university hospital, and all study participants.

## Authors’ contributions

All authors substantial contributions to the conception and design of the work; the acquisition, analysis, interpretation of data; have drafted the work and substantively revised it. All authors have approved the submitted version.

